# Cross-site imputation for recovering variables without individual pooled data

**DOI:** 10.1101/2024.12.19.24319364

**Authors:** Robert Thiesmeier, Paul Madley-Dowd, Nicola Orsini, Viktor H. Ahlqvist

## Abstract

In multi-site studies, it is common for some sites not to have recorded key variables. Although it is theoretically possible to use data from sites with recorded observations to impute the missing values, this process becomes challenging when data pooling is not feasible due to logistic or legal constraints. We therefore propose a multiple imputation approach — cross-site imputation — to recover any variables across sites without the need to pool individual-level data. The solution involves transporting predicted regression coefficients and variances from studies with observed data to impute missing variables at sites without data. The approach is illustrated in an applied example of recovering confounders across Swedish hospitals, and theoretical considerations are outlined. Given the increasing importance of multi-site studies in observational research, cross-site imputation could offer a practical approach for imputing variables that have not been recorded in some study sites.

## 1. Introduction

Collaborations both within and between countries have become the norm in modern observational research. These collaborative efforts may enhance precision and often allow for the study of more precisely defined exposures and outcomes [1, 2]. Additionally, they may improve the generalisability and transportability of findings, sometimes making the results more applicable to diverse populations [3]. However, due to regulatory constraints and the prioritization of timely results, a new standard has emerged wherein individual-level data is not pooled but qualitatively harmonized and analyzed at individual sites before being meta-analyzed [4]. This approach is sometimes referred to as ‘federated analysis’ [5], ‘common data models’ [2], or ‘distributed data network’ [6], but in principal, a similar analytical philosophy is applied.

Despite the benefits of these collaborative efforts, a common challenge is the inconsistent recording of variables across different sites [7]. Some sites may lack data on specific covariates of interest, whilst others have recorded them. A covariate that is 100% missing at one or more sites, i.e., not recorded, is commonly referred to as systematically missing data [8]. The challenge of systematically missing data becomes particularly important when such covariates are potential confounders that should be adjusted for in observational studies [9]. Currently, the default approach to dealing with this issue is:

i. excluding sites without the covariate data, which reduces statistical power and generalisability; or
ii. ignoring the missing covariate at certain sites and meta-analysing nonetheless, which can lead to biased inference due to lack of adjustment for confounders.

A less commonly employed alternative is to impute the missing values at sites where the covariate was not recorded by using information from the sites where data has been collected [10, 11, 12, 13, 14, 15]. However, when individual-level data cannot be pooled, standard imputation procedures are typically infeasible as they require all data to be at the disposal of the researcher [16, 17].

In this paper, we propose an imputation strategy that avoids the need to pool individual-level data by relying on sending model predictions across sites and using these to impute the missing values at sites that have not recorded the relevant covariate. We believe that this approach to recovering systematically missing data could be useful for international collaborations when important covariates, such as confounders, have not been recorded at all sites.

## 2. Methods

### 2.1. Missing data and multiple imputation

Missing data often requires careful considerations [18]. The default approach to handling missing values is often to exclude participants (or studies) with missing information, known as complete case analysis. While the simplicity of this approach is appealing, depending on the mechanism that underlies the missingness, it can lead to bias and loss of information [19, 20]. Several statistical methods have been proposed to resolve this problem, with multiple imputation being one of the most popular and flexible ones [21, 18]. In brief, missing values are imputed multiple times by drawing a value from the posterior distribution based on a model estimating the values for the missing variable(s) (the *imputation model*). For each draw of a value, an imputed data set is created. After the process has been repeated *M* times, an outcome model is fitted in each imputed data set. Estimates from all data sets are subsequently pooled using Rubin’s rules [21]. Tutorials for multiple imputation in clinical research have been published elsewhere [22]. However, existing multiple imputation techniques have been primarily developed for single-study settings with sporadically missing data, i.e., data in one or more variables is missing less than 100% [23]. In recent years, multiple imputation methods have been further extended and developed for individual-level multi-site pooling projects where researchers have all available data stored in one location [24, 13, 14, 15]. New methods are, however, needed to address the challenge of imputing key variables for sites with 100% missing data within multi-study collaborations under scenarios where individual-level data cannot be physically pooled due to practical limitations or regulatory restrictions.

### 2.2. Cross-site imputation

Typically, there is a wealth of information recorded at each site in a collaborative project with some variables having been recorded at all sites (e.g., age, disease status, sex, etc.). However, there are sometimes key variables, such as important confounders, that were never recorded at some sites. Imputing missing values without the need to transport the individual-level data requires sharing model parameters between studies. Unlike data, these model parameters are typically not under the same regulatory and practical constraints, as they are similar to sharing results for meta-analyses across studies [16]. Imputation across multiple sites has been previously explored in various forms [16, 17, 25, 26], and currently accommodates the imputation of continuous, binary, categorical variables [27]. We note that, under an assumption that the association between the auxiliaries and the imputation target are transferable across sites, cross-site imputation can provide an unbiased estimate [17, 26]. Although a strong assumption, it could also be noted that cross-site imputation can account for the distribution of auxiliary data at the site that needs imputation. In this paper, we illustrate this approach, discuss five practical steps for its implementation (Figure 1), and reflect on the required assumptions.

**Figure 1:**
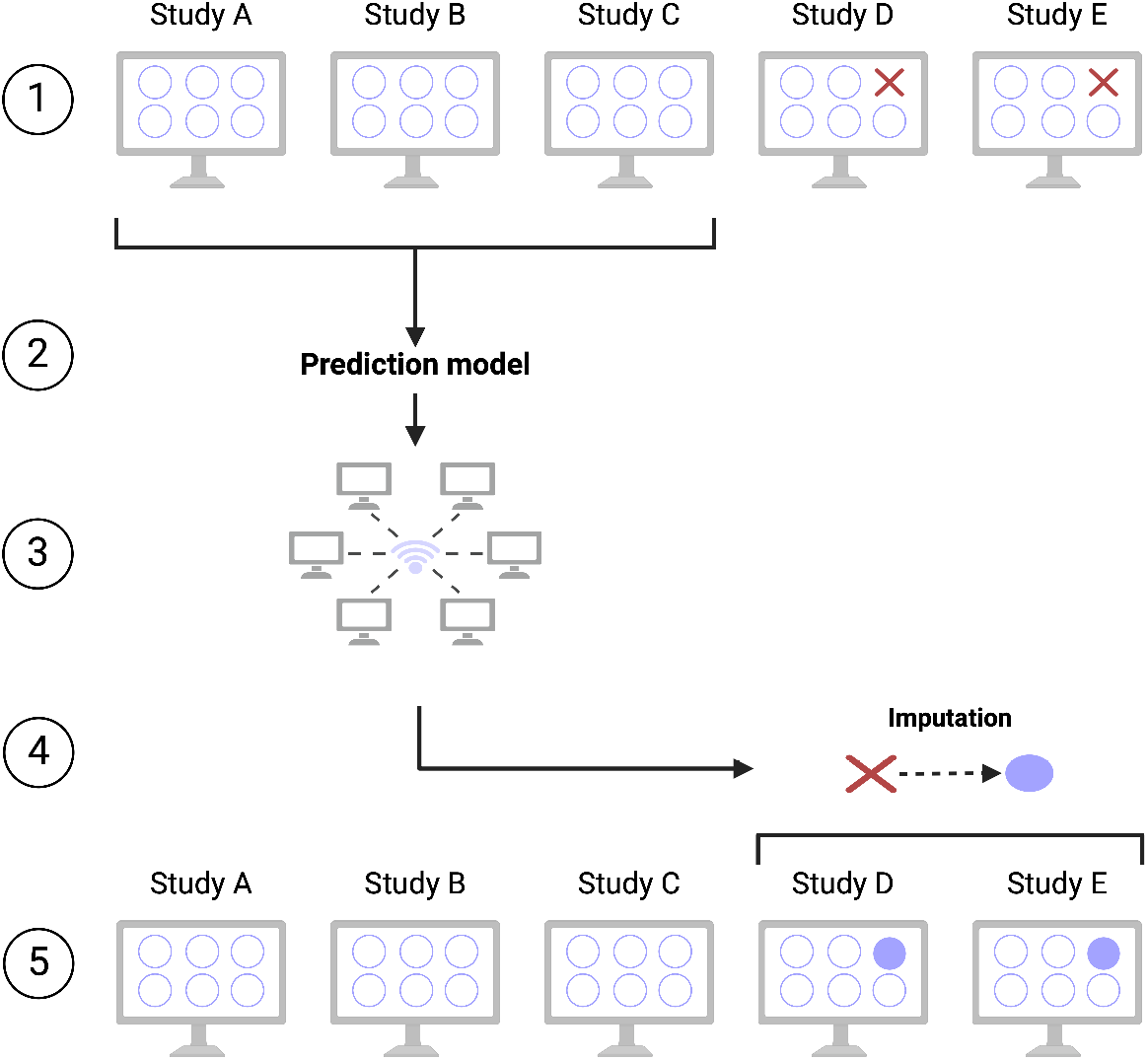
Visualisation of the cross-site imputation approach to recover variables in analyses with multiple studies when sharing and pooling individual-level data is not feasible. Each circle represents a variable in the data set whereas the cross represents a variable that has not been observed.

#### 1. Identify key variables that are not recorded at one or more sites

We first locate the studies with missing key variables. In turn, we can identify the sites with observed data on the missing variables that can be used for the imputation process in subsequent steps.

#### 2. Fit a prediction of the target variable at the sites with observed data

The next step is to fit a prediction model of the missing target variables identified in the previous step. The model typically includes a subset of variables that can best predict the missing values and should at least include all variables that are also considered in the outcome model. This includes the outcome itself and any transformation of variables (quadratic or interaction terms) [28, 29, 30]. Of note, the “imputation model” in this step is a prediction model of the conditional distribution of the target variable given covariates. Following the agreement on a suitable model, there are two options:

i. a single study is used to fit the prediction model; or
ii. multiple studies are used to fit the prediction model.

#### 3. Transfer prediction coefficients to sites with missing data

All sets of regression coefficients and their covariances from the prediction model(s) are shared with the site that requires imputation of missing values. If multiple sites were used to fit the prediction model, the site with missing data would receive multiple sets of prediction model parameters. A weighted average (e.g., inverse variance weighted) of all coefficients is then taken to proceed.

#### 4. Use predictions to impute the target variable

Before the imputation of missing values, we take a random draw from a normal distribution with the mean equal to the summary regression coefficient and a standard deviation equal to the corresponding standard error of the summary regression coefficient. In other words, we introduce some randomness in the model parameters that are used for the imputation to allow for uncertainty in the imputations [28]. Missing values are then imputed conditionally on the same set of covariates that have been used for the prediction model in Step 2. A technical overview of how imputations are simulated can be found in [27]. For each imputation, *M* imputed data sets are created and are analysed in the subsequent step. Currently, few studies have assessed the impact of the number of imputations for systematically missing data. In this paper, we use 20 imputations [29].

#### 5. Fit the outcome model and combine the estimates

The final step follows standard multiple imputation approaches. First, an outcome model is fitted to each imputed data set. Second, the resulting estimates are then combined with Rubin’s rules [21]. This process is done *only* at sites with missing data. The final estimates of the outcome model can then be used to proceed for the subsequent federated analysis.

### 3. Applied example: Multi-site study based on Swedish registers

To illustrate the cross-site imputation approach, we applied it to a multi-site study using Swedish registry data of children born between 1997 and 2014 from five major hospitals across Sweden. This hypothetical study aims to estimate the causal effect of maternal antidepressant use during pregnancy on the risk of neurodevelopmental conditions in offspring (autism, ADHD, or intellectual disability), a topic previously explored in the literature [31, 32, 33]. For this example, we assume that hospitals cannot share patient-level data directly. Available variables at all hospitals include the outcome — children’s diagnoses of neurodevelopmental conditions based on nationwide inpatient and outpatient records — and the exposure, which is maternal antidepressant use reported during antenatal visits or through pharmacy dispensations. Additionally, parental psychiatric history (a key confounder) is available at all sites, but for the purposes of this illustrative example, we drop this data from two hospitals. Though simplified, this scenario reflects real-world challenges in research collaborations, where some sites may lack key confounding variables. We implemented the following analyses in this hypothetical study:

#### (1) Approach 1 (unadjusted pooled estimate)

Federated analysis of all 5 studies *without* adjustment for parental psychiatric history;

#### (2) Approach 2 (adjusted complete case analysis)

Federated analysis using only the 3 studies with observed data on parental psychiatric history to be able to adjust for it, dropping the two studies that had systematically missing values;

#### (3) Approach 3 (adjusted for available data estimate)

Federated analysis using what is available at each site and pool estimates regardless (i.e., adjustment in only three studies in addition to the estimates from the other two studies without the adjustment for parental psychiatric history);

#### (4) Approach 4 & 5 (multiple study imputation adjusted pooled estimate)

Federated analysis after imputing missing values for parental psychiatric history at two hospitals. Two options were considered:

- A single study as the basis for imputation, or
- All studies as the basis for imputation.

#### (5) Approach 6 (full data estimate)

As a reference analysis where no variables are missing at any site, we implemented a federated analysis with adjustment for parental psychiatric history at all hospitals.

##### Fit the prediction model

In Hospitals 1 to 3 with observed data on the systematically missing variable - parental psychiatric history - we fit a logistic regression model on parental psychiatric history conditional on the main exposure - maternal antidepressants use - and the outcome - diagnosis of neurodevelopmental conditions in offspring. Once transferred to the sites with missing data, we performed 20 imputations at each site using these prediction models as the basis for the imputation. The analysis was performed with a recently developed software in Stata 18 [27]. A working example for this paper, providing data and computer code, can be found in the supporting files.

### 4. Applied results

The five hospitals were comparable in their distribution of patients with parental psychiatric history and diagnosis of a neurodevelopmental condition. Further, all hospitals had a similar distribution of maternal antidepressant use with an average of 2.6% users across hospitals (1). In the referent scenario (Approach 6), offspring exposed to maternal antidepressant use during pregnancy had 1.16 times the odds of being diagnosed with neurodevelopmental conditions compared to unexposed offspring (95% CI: 1.10, 1.23) with a standard error of 0.032. The effect estimate aligns with those previously reported in the literature [31, 32, 33], albeit more rigorous causal methods appear to not detect such associations [31, 33]. For illustrative purposes, data on parental psychiatric history in Hospital 4 and 5 were dropped for the subsequent analyses. Other variables in this study did not contain missing data.

First, in an analysis including all hospitals without adjusting for parental psychiatric history (Approach 1), the odds of neurodevelopmental conditions diagnosis were 1.76 times higher in offspring exposed to maternal antidepressant use during pregnancy compared to unexposed offspring. Second, restricting the analysis to hospitals 1-3 and adjusting for parental psychiatric history (Approach 2), the association attenuated to an odds ratio of 1.15. However, the standard error increased to 0.037 as a result of using fewer studies in the analysis. Third, when parental psychiatric history was adjusted for among hospitals with available data, and the hospitals without measurements were also included in the meta-analysis (Approach 3), the odds of neurodevelopmental conditions diagnosis were 1.29 times higher among children exposed to antidepressants. In Approach 4, using cross-site imputation with 20 imputations for the systematically missing parental psychiatric history from hospitals 4 and 5, we retained all hospitals in the analysis but with full adjustment for the covariate. The imputation was based on either a single hospital prediction model coefficients (Approach 4) or a weighted average of coefficients from three hospitals with complete data (Approach 5). Both of these approaches yielded similar results (OR 1.17), but with standard errors akin to that of the referent analysis where everything was measured.

**Table 1:** Descriptive information on the offspring participants of a study from five major hospitals in Sweden.

|  | **Hospital 1**<br>N=136,893 | **Hospital 2**<br>N=72,227 | **Hospital 3**<br>N=164,687 | **Hospital 4**<br>N=52,219 | **Hospital 5**<br>N=43,362 |
| --- | --- | --- | --- | --- | --- |
| **Maternal antidepressant use (%)** |  |  |  |  |  |
| No | 130,544<br>(97.7) | 68,637<br>(97.8) | 154,829<br>(97.1) | 49,255<br>(96.9) | 40,893<br>(97.5) |
| Yes | 3,091<br>(2.3) | 1,568<br>(2.2) | 4,590<br>(2.9) | 1,588<br>(3.1) | 1,028<br>(2.5) |
| **Parental history of psychiatric diagnosis (%)** |  |  |  |  |  |
| No | 90,226<br>(65.9) | 49,765<br>(68.9) | 116,276<br>(70.6) | 35,562<br>(68.1) | 29,395<br>(67.8) |
| Yes | 46,667<br>(34.1) | 22,462<br>(31.1) | 48,411<br>(29.4) | 16,657<br>(31.9) | 13,967<br>(32.2) |
| **Offspring diagnosis of a neurodevelopmental condition (%)** |  |  |  |  |  |
| No | 123,316<br>(90.1) | 67,983<br>(94.1) | 151,544<br>(92.0) | 47,902<br>(91.7) | 39,543<br>(91.2) |
| Yes | 13,577<br>(9.9) | 4,244<br>(5.9) | 13,143<br>(8.0) | 4,317<br>(8.3) | 3,819<br>(8.8) |

**Table 2:** The odds ratios and 95% confidence intervals of offspring diagnosis of neurodevelopmental conditions associated with maternal antidepressant use during pregnancy, derived from a federated analysis of five hospitals in Sweden. Data for parental psychiatric history were 100% missing at hospitals 4 and 5. OR = Odds Ratio; CI = Confidence Interval; SE = Standard Error.

| **Method** | **OR** | **95% CI** | **SE** |
| --- | --- | --- | --- |
| **Approach 1** (unadjusted pooled estimate): | 1.76 | 1.67, 1.86 | 0.049 |
| **Approach 2** (adjusted complete case analysis): | 1.15 | 1.08, 1.22 | 0.037 |
| **Approach 3** (adjusted for available data estimate) | 1.29 | 1.22, 1.36 | 0.036 |
| **Approach 4** (single study imputation adjusted pooled estimate) | 1.17 | 1.11, 1.24 | 0.033 |
| **Approach 5** (multiple study imputation adjusted pooled estimate) | 1.17 | 1.11, 1.24 | 0.033 |
| **Approach 6** (full data estimate) | 1.16 | 1.10, 1.23 | 0.032 |

#### Interpretation

In this case study using Swedish registry data, information on a key confounder — parental psychiatric history — was set to systematically missing in two hospitals. After applying cross-site imputation, we were able to recover this missing data and proceed with an adjusted federated analysis. The final estimates following imputation were closely aligned with those from the referent analysis, with similar standard errors. Notably, when parental psychiatric history was not accounted for in the two hospitals (Approach 3), the association appeared larger than expected. Furthermore, Approach 2 resulted in some loss of precision and, qualitatively, reduced generalisability by excluding data from these two hospitals.

### 5. Assumptions and considerations

Cross-site imputation may be a useful approach to impute variables that have not been recorded at some sites when pooling of individual-level data is not feasible. Previous simulation studies have demonstrated a promising performance of the approach, especially for continuous variables[17, 26]. However, certain qualitative considerations and assumptions may need to be highlighted.

First, the validity of cross-site imputation will depend on its central assumption: the association between the auxiliaries and the imputation target are transferable across sites. In other words, we assume that the relationship between the target variable and its predictors is equivalent across sites. Researchers are thus required to critically evaluate the transferability assumption. Important considerations include i) the structure and characteristics of the studies used to develop the prediction and fit the imputation (e.g., geographical proximity, similar participant characteristics, etc.), ii) whether there is a richer set of auxiliary variables recorded at some sites (e.g., ideal auxiliaries that are strong predictors of the missing variable) [34], and iii) if there are optimal measurements (’gold standard’ clinical measures) of the variables that have nor been recorded at some other site (e.g., comorbidities might be self-reported or clinically confirmed).

Second, researchers can either use a single study or multiple studies as the basis of imputation. To capture the heterogeneity between studies, considering multiple sites as the basis of imputation is in line with standard imputation approaches in collaborative projects where individual data can be pooled [15]. However, for cross-site imputation to be unbiased, one must assume that the transportability of auxiliary information holds across sites. We note that there can be situations where it may not be ideal to use data from multiple international studies for imputing missing values at one site. For example, if one or more sites within a larger collaboration are considered more similar to the site with missing data, restricting the data used for imputation to those sites could make the transportability assumption more plausible. In contrast, using all sites indiscriminately might violate this assumption, particularly if the more distal sites differ substantially from the target site. In addition, studies might have multiple variables with missing values. So far, this approach has been only considered for univariate imputation of continuous variables when data across studies cannot be shared [26]. An extension to a multivariate approach would need to take into account the joint distribution of missing variables conditional on a set of predictors that are observed in all study sites [17].

Last, practical considerations regarding the implementation of software to facilitate the imputation are described in detail elsewhere [27, 35]. Careful attention should be paid to the structure and naming of variables in the imputation model at all sites. This can be achieved with a commonly implemented master code or common data model shared between sites.

## 6. Discussion

In this paper, we proposed an approach for recovering missing variables in multi-site projects without the need to share individual-level data across studies. Despite many benefits of collaborative efforts in observational research, unrecorded or unobserved variables (systematically missing data) across different sites remain a major challenge. Current multiple imputation strategies require individual-level data from all studies to be in a single file. However, sharing individual-level data across sites is often not feasible due to logistic and legal constraints. Therefore, we proposed a cross-site imputation approach in which missing variables can be imputed without individual-level data from other studies. In a hypothetical multi-site study based on Swedish registry data from 5 major hospitals, we showed how the imputation approach can be used to effectively recover missing confounders and produce estimates that align with expectations (i.e., as if all data had been recorded). In the context of the recently proposed phases of methodological development [36], cross-site imputation is currently in phase II. Its present use, limitations, and further extension and potentials will be discussed in the following section.

Few studies have explored practical methods for imputing missing values without requiring individual-level data [17, 26, 16, 25, 37]. For instance, Secrest et al. [17] used a validation data set that contains complete observations of all variables to predict the target variable. The validation data site needs to share estimated parameters of the posterior prediction model which are in turn used to impute missing values at the target site. Alternatively, Li et al. [25] demonstrated how machine learning techniques can be applied to impute missing covariates in distributed data networks. Although their simulations yielded promising results, the study did not consider systematically missing data. Another approach, detailed by Cheng et al.[16], employed multiple imputation by chained equations for distributed health data networks. While the results were promising, the authors highlighted that the method’s lack of communication efficiency could limit its practical utility for researchers. In summary, while there has been previous work on imputing missing data without sharing individual-level data, we believe that cross-site imputation is flexible to use and a practically feasible method to retain any missing values in federated analyses without sharing individual data across sites. While we emphasized the method’s key assumption, *the transportability assumption*, and discussed practical considerations, further research and real-world applications are essential to fully evaluate its capabilities. Additionally, we have so far only considered univariate imputation of variables. A natural generalisation to the this approach can be a Fully Conditional Specification for mulitvariate imputation procedures. Further practical extensions could address situations where multiple variables are 100% missing or a mix of systematic and sporadic missingness occurs (e.g., one variable missing 100% and another variable missing at 50%). Of note, in this study, the study sites used to fit the prediction model had complete data.

## 7. Conclusion

In multi-site studies, logistic or legal barriers often prevent the pooling of individual-level data, rendering standard imputation methods infeasible, as these typically require all data to be accessible to the researcher. To address this challenge, we propose an approach—cross-site imputation—to recover missing variables without necessitating the pooling of individual-level data across sites. This method relies on the transfer of predicted imputation coefficients and their associated covariances derived from studies with observed data on the missing target variables, thereby eliminating the need for sharing individual-level data. Given the growing importance of multi-site studies in observational research, we believe that cross-site imputation could offer a valuable solution for addressing missing data that are not consistently recorded across all sites.

## Data Availability

Original data cannot be shared. working example is provided.

https://github.com/robertthiesmeier/cross_site_imputation.git

## 8. Data availability

Computer code used in this paper is illustrated and explained here https://github.com/robertthiesmeier/cross_site_imputation.git. Original data cannot be shared.

## 11. Additional information

All research was performed in accordance with relevant guidelines/regulations. The Swedish Ethical Review Authority (2020-05516) approved the study. The figure was created with BioRender.com.

